# Effectiveness and Safety of MSC Cell Therapies for Hospitalized Patients with COVID-19: A Systematic Review and Meta-analysis

**DOI:** 10.1101/2021.10.05.21264559

**Authors:** Wenchun Qu, Zhen Wang, Erica Engelberg-Cook, Abu Bakar Siddik, Guojun Bu, Julie G. Allickson, Eva Kubrova, Arnold I. Caplan, Joshua M. Hare, Camillo Ricordi, Carl J. Pepine, Joanne Kurtzberg, Jorge M. Pascual, Jorge M. Mallea, Ricardo L. Rodriguez, Tarek Nayfeh, Samer Saadi, Elaine M. Richards, Keith March, Fred P. Sanfilippo

**Affiliations:** Department of Pain Medicine, Mayo Clinic, Jacksonville, Florida; Center for Regenerative Medicine, Mayo Clinic, Jacksonville, Florida; Evidence-Based Practice Center, Mayo Clinic, Rochester, Minnesota; Robert D. and Patricia E. Kern Center for the Science of Health Care Delivery, Mayo Clinic, Rochester, Minnesota; Department of Neuroscience, Mayo Clinic, Jacksonville, Florida; Center for Regenerative Medicine, Mayo Clinic, Rochester, Minnesota; Department of Physical Medicine and Rehabilitation, Mayo Clinic, Rochester, Minnesota; Skeletal Research Center, Biology Department, Case Western Reserve University, Cleveland, Ohio; Interdisciplinary Stem Cell Institute and Cardiology Division, Department of Medicine, University of Miami, Miller School of Medicine, Miami, Florida; Department of Surgery, Diabetes Research Institute and Cell Transplant Center, University of Miami Miller School of Medicine; Division of Cardiovascular Medicine, and Center for Regenerative Medicine, University of Florida, Gainesville, Florida; Marcus Center for Cellular Cures, Duke University School of Medicine; Division of Pulmonary, Allergy and Sleep Medicine, Department of Medicine, Mayo Clinic, Jacksonville, Florida; Co-Chair Regenerative Medicine Committee, American Society Plastic Surgeons; Department of Physiology and Functional Genomics, Center of Regenerative Medicine, University of Florida, Gainesville, Florida; Division of Cardiovascular Medicine, Center for Regenerative Medicine, University of Florida, Gainesville, Florida; Department of Pathology and Laboratory Medicine, Department of Health Policy and Management, Emory University, Atlanta, Georgia

**Keywords:** MSC, COVID, Cytokine storm, Pulmonary, Hospital recovery

## Abstract

MSC (a.k.a. mesenchymal stem cell or medicinal signaling cell) cell therapies have shown promise in decreasing mortality in ARDS and suggest benefits in treatment of COVID-19 related ARDS. We performed a meta-analysis of published trials assessing the effectiveness and adverse events (AE) of MSC cell therapy in individuals hospitalized for COVID-19. Systematic searches were performed in multiple databases through April 8^th^, 2021. Reports in all languages including randomized clinical trials (RCTs), comparative observational studies, and case series/case reports were included. Random effects model was used to pool outcomes from RCTs and comparative observational studies. Outcome measures included all-cause mortality, serious adverse events (SAEs), AEs, pulmonary function, laboratory and imaging findings. A total of 413 patients were identified from 25 studies, which included 8 controlled trials (3 RCTs), 5 comparative observational studies, (n=300) and 17 case-series/case reports (n=113). The patients age was 60.5 years (mean), 33.7% were women. When compared with the control group, MSC cell therapy was associated with reduction in all-cause mortality (RR=0.31, 95% CI: 0.12 to 0.75, I^2^=0.0%; 3 RCTs and 5 comparative observational studies, 300 patients), reduction in SAEs (IRR=0.36, 95% CI: 0.14 to 0.90, I^2^=0.0%; 3 RCTs and 2 comparative studies, n=219), no significant difference in AE rate. A sub-group with pulmonary function studies suggested improvement in patients receiving MSC. These findings support the potential for MSC cell therapy to decrease all-cause mortality, reduce SAEs, and improve pulmonary function compared to conventional care. Large scale double-blinded, well-powered RCTs should be conducted to further explore these results.

## INTRODUCTION

As of August 2021, the pandemic caused by COVID-19 (SARS-CoV-2) has infected more than 200 million and contributed to the death of more than 4 million individuals worldwide.^1^ While the rapid development of effective vaccines is helping to mitigate this pandemic, there is a rising incidence of infection with the more highly transmissible Delta variant that is expected to increase COVID related diseases, and is capable of breakthrough disease in vaccinated individuals.^2^ To date, most (81%) infected patients have a mild to moderate disease course and recover within two or three weeks.^3^ However, among patients requiring hospitalization the mortality ranges from 5 to 17%^4, 5^ and among those requiring ICU admission, range from 26 to 78%.^6,7^Additionally, emerging new variants of the virus have raised concerns about vaccine efficacy^2, 8^, virus transmissibility, susceptibility and disease severity.^9^ Although viral mutation occurs in a natural cycle, often including removal of lethal or deleterious variants from the pool, new variants will likely extend the duration of the pandemic.^9^

In severe cases, SARS-CoV-2 leads to fatal acute respiratory distress syndrome (ARDS) with diffuse alveolar damage and cellular fibromyxoid exudate associated with monocyte and macrophage infiltration.^10, 11^ Respiratory distress typically arises 7 to 10 days post-symptom onset, with manifestations of immune dysregulation including cytokine release syndrome [IL-1, IL-6, IL-8, IL2R, IL-10, and tumor necrosis factor-α (TNF-α)], also known as “cytokine storm”, lymphopenia (CD4+ and CD8+ T cells), and decreased interferon-γ (INF-γ) expression in CD4+ T cells.^11, 12^ The inverse correlation between cytokine storm and lower CD4+ and CD8+ counts suggests that the cytokine response may dampen adaptive immunity.^13^

Treatments for patients with respiratory complications from COVID infection have evolved since the onset of the pandemic. These have focused on supportive approaches including mechanical ventilation, high-flow nasal oxygen, and ECMO, convalescent plasma, as well as anti-inflammatory and immuno-modulatory therapies ranging from corticosteroids to monoclonal antibodies targeting specific cytokines such as IL-6.^14, 15^ While several drug therapies have suggested promising results^14, 15^, their potential benefits must be balanced with concerns about side effects and secondary infections.^14^ New treatments focused on mitigating the underlying cellular and molecular mediators of diseases are urgently needed to address the pathophysiological processes that lead to death or long-term sequelae.

MSC cell therapies have shown promise in modulating responses in inflammatory diseases.^16–18^ Previously known as “mesenchymal stem cells” or “mesenchymal stromal cells” and more recently as “medicinal signaling cells,”^19^ MSCs are small spindle shaped cells found in the extracellular matrix and are most commonly sourced from bone marrow, adipose tissue, and umbilical cord and they can also be isolated from other tissues.^20^ They express the cell surface markers CD44, CD90, CD105 but do not express CD34, CD45 or HLA-DR.^21^ MSCs home to sites of injury and inflammation where they exert immunomodulatory effects, largely via a paracrine mode of action.^22, 23^ Their immunomodulatory activity arises through several well characterized effects that include: (a) suppression of proliferation and action of B cells, T cells, natural killer cells, and dendritic cells; (b) polarization of monocytes to anti-inflammatory M2 macrophages; (c) differentiation of T effector cells towards Treg cells; and (d) modulation of cytokine secretion towards increased production of IL-4, IL-10 with reduced production of TNF-α, IL-12 and IL-6.^16, 21, 24, 25^ MSCs may also enhance tissue repair and regeneration by secretion of factors that limit apoptosis and foster endogenous progenitor cell cycling.^26, 27^

With regard to the lung, MSC cell therapy has been shown to repair lung epithelium by increasing alveolar ATP, transferring mitochondria to the damaged alveolar epithelium^28^, maintaining balance in the renin-angiotensin system, and improving endogenous repair of pulmonary endothelial cells by enhancing their microenvironment. They also improve the alveolar-capillary barrier function of the lung^22, 29, 30^. Studies of COVID-19 patients suggest that MSC cell therapy can reduce inflammation by ameliorating anti-inflammatory and trophic factor expression with notable decreases in C-reactive protein (CRP), TNF-α, and cytokine-secreting immune cells.^31, 32^ In addition, MSCs have powerful antifibrotic effects and may alleviate pulmonary fibrosis.^33^ Finally, MSCs demonstrate antimicrobial activity via enhanced production of peptide cathelicidin/LL-37 production^34^, which also has antiviral activity.^35^ Preclinical study suggests that MSCs have antimicrobial properties *in vivo* and thus may limit microbial superinfections in the context of viral infections.^36^

Clinically, MSC cell therapy has been studied in viral and non-viral-induced ARDS, and appears to reduce mortality.^16^ Importantly, early data from the pandemic suggested that infusion of various MSC preparations could potentially reduce COVID-19 mortality and morbidity^37^. However, these initial clinical reports had small patient numbers and limited outcome measures. To address this issue, we conducted a systematic review of the current literature on safety, efficacy, and cytokine responses to MSC cell therapy in patients with COVID-19 infections.

## METHODS

### Search strategy

An extensive search of databases from the database inception to April 8^th^, 2021 in all languages was performed. Databases included EBM Reviews – Cochrane Central Register of Controlled Trials, EBM Reviews – Cochrane Database of Systematic Reviews, Ovid MEDLINE, Epub Ahead of Print, In-Process, In-Data-Review & Other Non-Indexed Citations and Scopus. A medical reference librarian developed and implemented the search strategy, with feedback from the investigators. Controlled vocabulary supplemented with keywords were used to search for MSC cell therapy for patients with COVID-19 or SARS-CoV-2 infection. The exact strategy is available in Appendix A.

### Eligibility criteria

We included all published randomized clinical trials, comparative observational studies and case series/case reports that evaluated the safety and/or effectiveness of stem cells administered to patients hospitalized with COVID-19 or SARS-CoV-2 infection. Studies were excluded if original data were not reported (such as a narrative review, editorials, or erratum). The detailed inclusion and exclusion criteria were listed in Appendix Table A1.

### Outcome measures

The outcomes of interest included all-cause mortality, serious adverse events (SAEs) and mild adverse events (AEs). SAEs and AEs were defined according to the original studies or CBER criteria^38^. Mortality was reported as a separate outcome and not included in SAEs in this review.

Other evaluated outcomes included pulmonary functional measures [e.g., PaO_2_/ FiO_2_ ratio and oxygenation index; laboratory measures including lymphocyte count, D-dimer, procalcitonin (PCT), CRP, IL-6; and imaging findings including computed tomographic (CT) of the lung].

### Study selection process

Independent reviewers, working in pairs, screened the titles and abstracts of all citations. Studies included by either reviewer were retrieved for full-text screening. Independent reviewers, again working in pairs, screened the full-text version of eligible studies. Conflicts between the reviewers were resolved by a third senior investigator (Figure 1).

**FIGURE 1:**
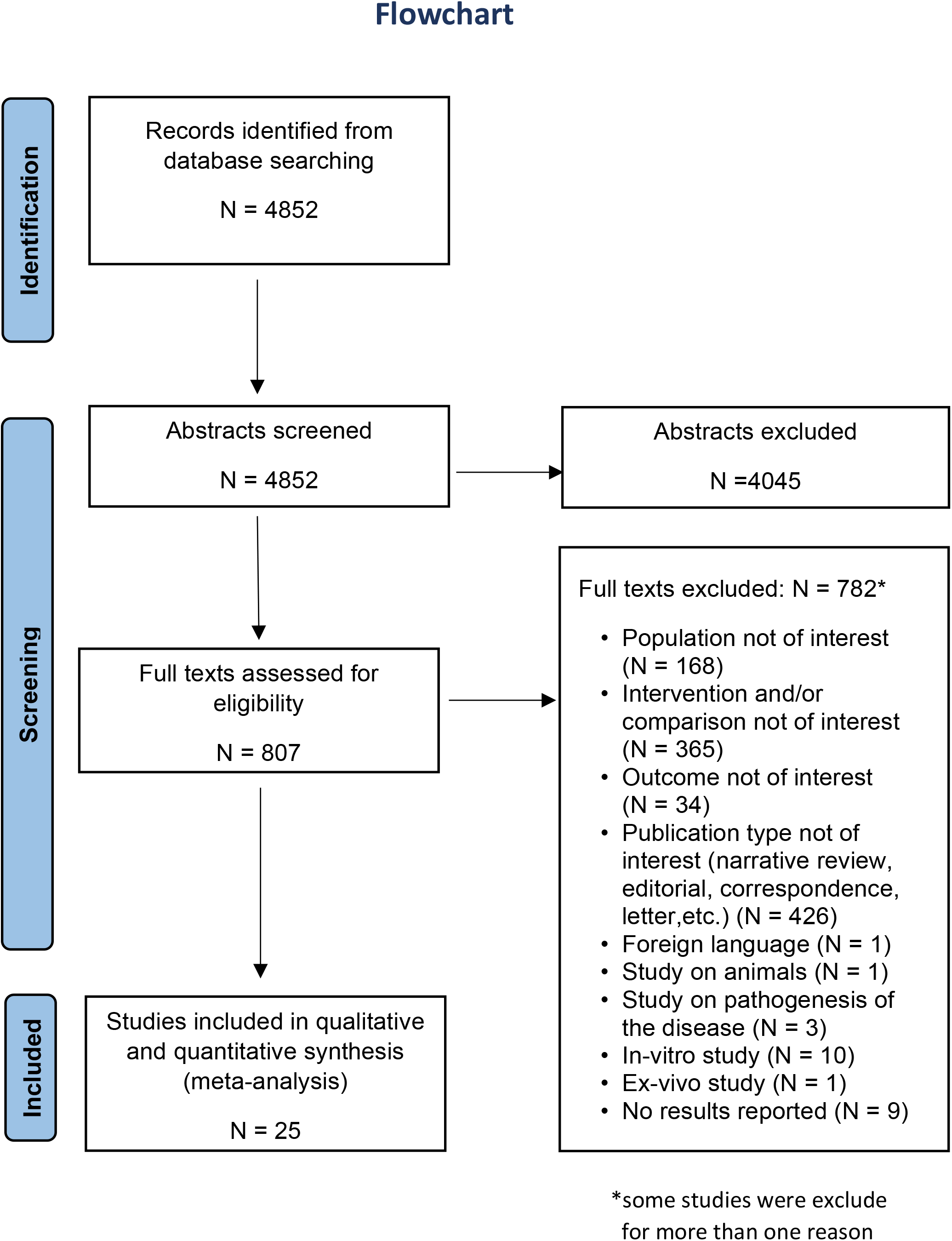
PRISMA 2009 flow diagram.

### Data collection

A pilot-tested standardized data extraction form was used to extract data. Independent reviewers extracted study details. An additional reviewer reviewed data extraction and resolved conflicts.

### Risk of bias

We used the Cochrane Collaboration’s Risk of Bias 2 tool to assess risk of bias for RCTs^39^, the Newcastle-Ottawa tool for comparative observational studies,^40^ and the risk of bias tool developed by Murad et.al for case series/case report.^41^

### Statistical analysis

To evaluate the comparative effectiveness of MSC cell therapy versus control groups, we calculated relative risk (RR) for mortality and incidence rate ratio (IRR) for SAEs and AEs from RCTs and comparative observational studies. The DerSimonian-Laird random-effects model with Hartung-Knapp-Sidik-Jonkman variance correction was used to combine effect size from included studies.^42^ Treatment group continuity corrections were used to adjust double-zero-event studies (i.e., 0 event in both groups).^43^ Heterogeneity between studies was evaluated with the *I*^2^ indicator. Two-tailed p value<0.05 was considered as statistically significant. All statistical analyses were conducted using Stata version 17 (StataCorp LLC, College Station, TX).

## RESULTS

The literature search identified 4852 distinct citations. Twenty-five studies with a total of 413 unique patients met the inclusion and exclusion criteria and were included in the analyses as shown in Figure 1.

Of the 25 included studies, we included 3 RCTs,^37, 44, 45^ 5 comparative observational studies,^46–50^ and 17 case-series/case reports.^51–67^ The studies were conducted in seven countries, including Asia (n=16), Europe (n=6), North America (n=2), and South America (n=1).

The overall average age of the participants was 60.5 years (range: 19 to 83 years), with 139 (33.7%) female. Among the patients, 133 (32.2%) were deemed as “critically ill”, 268 (64.9%) “severely ill”, and only 12 (2.9%) “moderately ill”. Hypertension, diabetes mellitus and obesity were the most 3 common comorbidities (Table 1). The follow-up durations were reported in 21 studies (n=333) ranging between 4 days to 196 days after the onset of treatment.

**TABLE 1:** Patient and product characteristics.

| Publication author, year | Country | Study Type | No. of Patients (Treatment; Control) | Age, median (IQR)/mean (SD), year | Female No. (%) | Comorbidities No. (%) | COVID-19 Severity: No. (%) | MSC Source | Dose [MSCs]* * | Administration Route | Frequency (schedule) |
| --- | --- | --- | --- | --- | --- | --- | --- | --- | --- | --- | --- |
| Shi et al, 2021<sup>45</sup> | China | RCT | 100 (65; 35) | Treatment: Mean: 60.7 (9.1)<br>Control: Mean: 59.9 (7.7) | Treatment 28(43.1)<br>Control 16(45.7) | Treatment HTN: 17(26.2)<br>DM: 12(18.5)<br>COPD: 4(6.2)<br>Control HTN: 10(28.5)<br>DM: 5(14.28)<br>COPD: 3(8.5) | Treatment Severe: 65(100)<br>Control Severe: 35(100) | Wharton-Jelly | 4 x 10E7 per dose | IV | 3 (D0, D3, D6) |
| Lanzoni et al, 2021<sup>44</sup> | USA | RCT | 24 (12; 12) | Treatment: Mean: 58.5 (15.9)<br>Control Mean: 58.8(11.6) | Treatment 7(58.3)<br>Control 4(33.3) | Treatment HTN: 7(58.3)<br>DM: 5(41.6)<br>CAD: 1(8.3)<br>Obesity: 11(91.6)<br>Control HTN: 9(75)<br>DM: 6(50)<br>CAD: 3(25)<br>Obesity: 5(41.6)<br>Cancer: 1(8.3) | Treatment Severe: 12(100)<br>Control Severe: 12(100) | Umbilical cord | 100±20 x 10E6 per dose | IV | 2 (D0, D3) |
| Shu et al, 2020<sup>37</sup> | China | RCT | 41 (12; 29) | Treatment: Mean: 61 (17.8)<br>Control Mean: 57.8 (15.7) | Treatment 4(33.3)<br>Control 13(44.8) | Treatment HTN: 3(25)<br>DM: 3(25)<br>Control HTN: 6(20.6)<br>DM: 5(17.2) | Treatment Severe: 12(100)<br>Control Severe: 29(100) | Umbilical cord | 2 x 10E6 per kg body wt. | IV | 1 |
| Xu et al, 2021<sup>49</sup> | China | COS | 44 (26; 18) | Treatment Mean: 58.3 (12.4) | Treatment 9(50) | NR; NR | Treatment | Menstrual blood | 3 x 10E7 | IV | 3 (D0, D2, D4) |
|  |  |  |  | Control<br>Mean: 61.1<br>(11) | Control<br>5(27.7) |  | Critically<br>ill:<br>10(38.4)<br>Severe:<br>16(61.5)<br>Control<br>Critically<br>ill: 8(44.4)<br>Severe:<br>10(55.5) |  | per<br>dose |  |  |
| Leng et al,<br>2020<sup>47</sup> | China | COS | 10 (7; 3) | Treatment<br>Mean: 57<br>(7.4)<br>Control<br>Mean: 65<br>(16.4) | Treatment<br>3(42.8)<br>Control<br>3(100) | Treatment<br>HTN: 1(14.2) | Treatment<br>Critically<br>ill:10<br>(14.2)<br>Severe:<br>4(57.1)<br>Moderate:<br>2(28.5)<br>Control:<br>Severe:<br>3(100) | NR | 1 x 10E6<br>per kg<br>body<br>wt. | IV | 1 |
| Meng et al,<br>2020<sup>48</sup> | China | COS | 18 (9; 9) | Treatment<br>Mean: 45.1<br>(10.1)<br>Control<br>Mean: 49.5<br>(10.6) | Treatment<br>2(22.2)<br>Control<br>5(55.5) | Treatment<br>HTN: 2(22.2)<br>DM: 1(11.1)<br>FLD: 1(11.1)<br>Control<br>HTN: 1(11.1)<br>BA: 1(11.1) | Treatment<br>Severe:<br>4(44.4)<br>Moderate:<br>5(54.3)<br>Control:<br>Severe:<br>4(44.4)<br>Moderate:<br>5(54.3)<sup>#</sup> | Umbilical<br>cord | 3 ×<br>10E7<br>per<br>dose | IV | 3 (D0, D3, D6) |
| Haberle et<br>al, 2021<sup>46</sup> | German<br>y | COS | 23 (5;<br>18) | Treatment<br>Median: 39<br>(32-50)<br>Control<br>Median: 59<br>(54-79) | Treatment<br>2(40)<br>Control<br>5(27.7) | Treatment<br>HTN: 1(20)<br>Control<br>HTN: 13(72.2)<br>DM: 2(11.1)<br>CAD: 2(11.1)<br>COPD: 1(5.5) | Treatment<br>Critically<br>ill: 5(100)<br>Control:<br>Critically<br>ill: 18(100) | NR | 1 x 10E6<br>per kg<br>body<br>wt. | IV | Cohort A-3<br>(NR), Cohort<br>B-2 (NR)<br>[Cohort A= 2<br>pts, Cohort<br>B= 3 pts] |
|  |  |  |  |  |  | AF: 2(11.1)<br>BA: 1(5.5)<br>Smoking:<br>3(16.6) |  |  |  |  |  |
| Singh et al, 2020<sup>50</sup> | USA | COS | 40(6; 34) | Treatment Mean: 56.3 (19.9)<br>Control Mean: 66.8 (13.6) | Treatment 1(16.6)<br>Control 8(23.5) | Treatment HTN: 3(50)<br>DM: 2(33.3)<br>HL: 3(50)<br>Obesity: 3(50)<br>CKD: 1(16.6)<br>AF: 16.6%<br>Osteoporosis: 1(16.6)<br>COPD/ BA: 1(16.6)<br>Control HTN: 18(52.9)<br>DM: 8(23.5)<br>Obesity: 6(17.6)<br>COPD/ BA: 7(20.5) | Treatment Critically ill: 6(100)<br>Control Critically ill 34(100) | Heart tissue* | 15 x 10E7 per dose | IV | Cohort A- 1<br>Cohort B- 2 (D0, D7)<br>[Cohort A= 4 pts, Cohort B= 2 pts] |
| Chen et al, 2020<sup>51</sup> | China | CS | 25 (25; - ) | Median: 70 (59-71) | 5(20) | NR | Severe: 25(100) | NR | 1 x 10E6 per kg body wt. | IV | Cohort A- 1,<br>Cohort B- 2 (D0, D5),<br>Cohort C- 3 (D0, D5, D10)<br>[Cohort A= 7 pts, Cohort B=7 pts, Cohort C= 11 pts] |
| Guo et al, 2020<sup>53</sup> | China | CS | 31 (31; - ) | Median: 70 (62-71) | 6(19.3) | HTN: 13(41.9)<br>COPD: 6(19.3)<br>DM: 5(16.1)<br>CAD: 5(16.1) | Critically ill: 8(25.9)<br>Severe: 23(74.1)<sup>#</sup> | Umbilical cord | 1 x 10E6 per kg body wt. | IV | Cohort A- 1<br>Cohort B- 2 (NR),<br>Cohort C- 3 (NR) |
|  |  |  |  |  |  |  |  |  |  |  | [Cohort A= 11 pts, Cohort B=9 pts, Cohort C= 11 pts] |
| Peng et al, 2020<sup>58</sup> | China | CR | 1 (1; -) | Mean: 66 | 1(100) | 0(0) | Severe: 1(100) | Umbilical cord | 1 x 10E6 per kg body wt. | IV | 3 (D0, D3, D6) |
| Zhang et al, 2020<sup>67</sup> | China | CR | 1 (1; -) | Mean: 54 | 0(0) | DM: 1(100) | Critically ill: 1(100) | Wharton-Jelly | 1 x 10E6 per kg body wt. | IV | 1 |
| Tang et al, 2020<sup>61</sup> | China | CR | 2 (2; -) | Mean: 54 (24) | 1(50) | HTN: 1(50)<br>CAD: 1(50) | Severe: 2(100) | Menstrual blood | 1 x 10E6 per kg body wt. | IV | 3 (D0, D1, D3) |
| Zhu et al, 2020<sup>65</sup> | China | CR | 1 (1; -) | Mean: 48 | 0(0) | 0(0) | Critically ill: 1(100) | Umbilical cord | 1 x 10E6 per kg body wt | IV | 1 |
| Zengin et al, 2020<sup>64</sup> | Turkey | CR | 1 (1; -) | Mean: 72 | 0(0) | HTN: 1(100)<br>DM: 1(100)<br>HL: 1(100) | Critically ill: 1(100) | Umbilical cord | 0.7 × 10E6 per kg body wt & 0.3 × 10E6 per kg body wt | IV & Intratracheal | 2 (D0, D5) |
| Yilmaz R et al, 2020<sup>63</sup> | Turkey | CR | 1 (1; -) | Mean: 51 | 0(0) | 0(0) | Critically ill: 1(100) | Wharton-Jelly | 3 × 10E6 per kg body wt. (1<sup>st</sup> - 3<sup>rd</sup> dose), 2 × 10E6 per kg body wt. & 1<br>× 10E6<br>per kg<br>body<br>wt. (4th<br>dose) | IV (1<sup>st</sup> - 3<sup>rd</sup> dose) IV & Intrathecal (4<sup>th</sup> dose) | 4 (D0, D3, D6, D9) |
| Tao et al,<br>2020<sup>62</sup> | China | CR | 1 (1; -) | Mean: 72 | 0(0) | HTN: 1(100)<br>DM: 1(100) | Critically<br>ill: 1(100) | Wharton<br>-Jelly | 1.5 x<br>10E6<br>per kg<br>body wt | IV | 5 (D0, D2, D4,<br>D6, D8) |
| Feng et al,<br>2020<sup>52</sup> | China | CS | 16 (16; -<br>) | Mean: 61.7<br>(10) | 4(25) | HTN: 8(50)<br>DM: 6(37.5)<br>CKD: 3(18.7)<br>Anemia:<br>1(6.25)<br>BA: 1(6.25)<br>HBV: 1(6.25) | Critically ill<br>7(43.7)<br>Severe<br>9(56.3) | Umbilical<br>cord | 1 ×<br>10E8<br>per<br>dose | IV | 4 (D0, D2, D4,<br>D6) |
| Liang et al,<br>2020<sup>56</sup> | China | CR | 1 (1; -) | Mean: 65 | 1(100) | HTN: 1(100)<br>DM: 1(100)<br>Anemia: 1(100) | Critically<br>ill: 1(100) | Umbilical<br>cord | 5 ×<br>10E7<br>per<br>dose | IV | 3 (D0, D3, D6) |
| Rich et al,<br>2020<sup>66</sup> | Spain | CR | 1 (1; -) | NR | NR | NR | Severe;<br>1(100) | Bone<br>marrow | 1 x 10E6<br>per kg<br>body wt | IV | 1 |
| Sahin et al,<br>2021<sup>59</sup> | Turkey | CR | 1 (1; -) | Mean: 33 | 1(100) | NR | Critically<br>ill: 1(100) | NR | NR | IV | 2 (D0, D3) |
| Lu et al,<br>2021<sup>57</sup> | China | CR | 1 (1; -) | Mean: 32 | 0(0) | 0(0) | Severe:<br>1(100) | Menstru<br>al blood | 3000U<br>(1<sup>st</sup> &<br>3<sup>rd</sup><br>dose,<br>2000U<br>(2<sup>nd</sup><br>dose) | IV | 3 (D0, D1, D3) |
| Iglesias et<br>al, 2021<sup>55</sup> | Mexico | CS | 5 (5; -) | Mean: 52.6<br>(13.5) | 1(20) | HTN: 1(20)<br>DM: 2(40)<br>HL: 1(20)<br>Obesity: 3(60) | Critically<br>ill: 5(100) | Umbilical<br>cord | 1 x 10E6<br>per kg<br>body wt | IV | 1 |
|  |  |  |  |  |  | Hypothyroidism: 1(20)<br>PAD: 1(20)<br>PF: 1(20) |  |  |  |  |  |
| Hashemian et al 2021<sup>54</sup> | Iran | CS | 11 (11; -) | Mean: 53.8 (10.3) | 3(27.2) | HTN: 3(27.2)<br>DM: 4(36.3)<br>Cardiomyopathy: 1(9.1)<br>HD/CLL: 1(9.1) | Critically ill: 11(100) | Umbilical cord (6 pts) or Placenta (5 pts) | 2 x 10E8 per dose | IV | 3 (D0, D2, D4) |
| Sanchez-Guijo et al, 2020<sup>60</sup> | Spain | CS | 13 (13; -) | Mean: 60.3 (7.8) | 1(7.6) | HTN: 5(38.4)<br>COPD: 2(15.3)<br>Obesity: 2(15.3)<br>DM: 1(7.6)<br>HTD: 1(7.6)<br>HBV: 1(7.6)<br>Hypothyroidism: 1(7.6) | Critically ill: 13 (100) | Adipose tissue | 0.98 x 10E6 per kg body wt | IV | Cohort A- 1<br>Cohort B- 3 (D0, D2, D4)<br>Cohort C- 2 (D0, D2/3)<br>[Cohort A= 2 pts, Cohort B= 1 pt, Cohort C= 10 pts] |

Abbreviations: RCT, randomized control trial; COS, comparative observational study; CS, case series; CR, case report; HTN, hypertension; CAD, coronary artery disease; CKD, chronic kidney disease; COPD, chronic obstructive pulmonary disease; DM, diabetes mellitus; HBV, hepatitis B virus; HL, hyperlipidemia; HD, hodgkin's disease; CLL, chronic lymphocytic lymphoma; AF, atrial fibrillation; FLD, fatty liver disease; BA, bronchial asthma; CHF, congestive heart failure; HTD, hyperthyroidism; PAD, pulmonary artery disease; PF, pulmonary fibrosis; NA, not applicable; NR, not reported; F/U, follow-up; SD, standard deviation; IQR, interquartile range; D, Day

The overall risk of bias was deemed low for RCTs and high for comparable and single-arm observational studies (Appendix Table A2-4).

### Intervention characteristics

Allogeneic culture-expanded MSCs were used in all 25 studies (n= 255 patients), of which MSCs were derived from umbilical cord in 14 studies (n=157), menstrual blood in 3 studies (n=29), bone marrow in 1 study (n=1), adipose tissue in 1 study (n=13), heart tissue in 1 study (n=6), combined placenta (n=5)and umbilical cord (n= 6)in 1 study, and non-specified sources in 4 studies (n=38). MSCs were administered via the systemic intravenous (IV) route in 23 studies (n=253), combined IV and intratracheal routes in 1 study (n=1), and combined IV and intrathecal routes in 1 study (n=1). The dose of MSCs was weight-adjusted ranging from 1 to 3 million cells per kilogram body weight in 15 studies (n=107). Eight studies (n=146) used a fixed dose between 30 million to 200 million MSCs. One study (n=1) used 2000U and 3000U MSCs, with undefined units.^57^ One study (n=1) did not report the cell dose.^59^ Frequency of the doses used are between 1 to 5 (Table 1). Patients in treatment groups received MSC cell therapy in addition to institutionally defined standard of care while patients in reference groups received institutionally defined standard of care only.

### Mortality

When compared to the control group, MSC cell therapy administration was associated with reduced mortality (RR=0.31, 95% CI: 0.12 to 0.75, I^2^=0.0%; 3 RCTs and 5 comparative observational studies, 300 patients; Figure 2). In all 25 studies including case series and case reports, 21 patients (8.2%) who received MSC cell therapy died.

**FIGURE 2:**
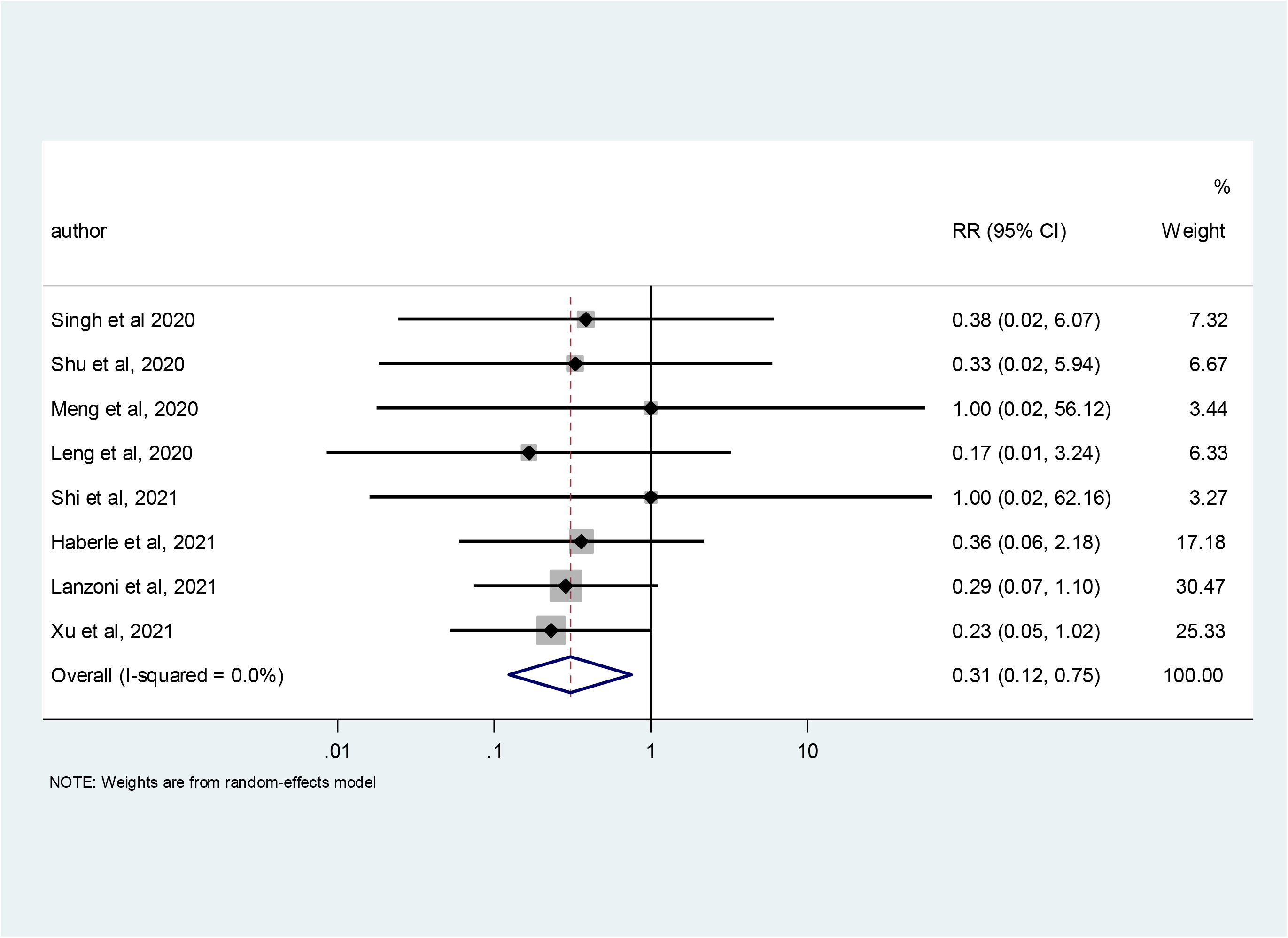
Mortality.

### Serious Adverse Events

MSC cell administration was associated with fewer SAEs compared to control group (IRR=0.36, 95% CI: 0.14 to 0.90, I^2^=0.0%; 3 RCTs and 2 comparative observational studies, 219 patients; Figure 3). Of all 24 studies that reported SAEs, a total of 33 incidences of SAEs were reported in 250 patients who received MSC cell therapy.

**FIGURE 3:**
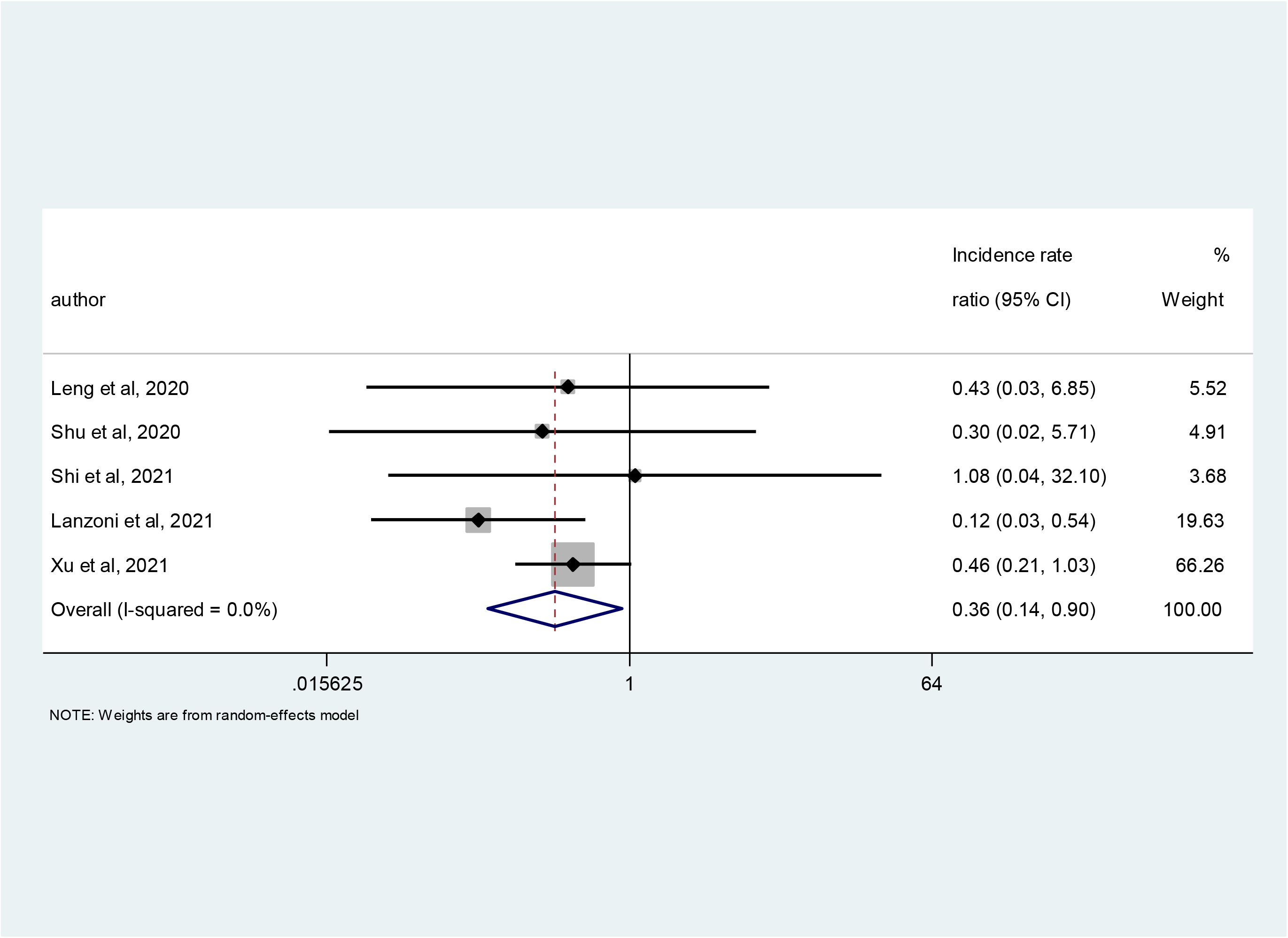
Serious adverse events.

### Adverse Events

No significant difference in AEs was found between MSC cell administration group and control group (IRR=0.86, 95% CI: 0.53 to 1.38, I^2^=32.6%; 3 RCTs and 2 comparative observational studies, 219 patients; Figure A1). Of all 23 studies that reported AEs, a total of 172 incidences of AEs were reported in 234 patients who received MSC cell therapy.

### Pulmonary and systemic changes

Longitudinal assessments of PaO_2_ to FiO_2_ ratios before and after MSC cell therapy was reported in 12 patients with MSC cell therapy, of whom 8 (66.7%) showed increased PaO_2_/FiO_2_, 4 (33.3%) patients showed decreased PaO_2_/FiO_2_ (Table 2).

**TABLE 2:** Pulmonary, laboratory and imaging outcomes.

| **Author, year** | **Pulmonary & systemic outcomes** | **Inflammatory biomarkers and leukocyte count** | **CT imaging** | **F/U (No. of days after 1st dose)** |
| --- | --- | --- | --- | --- |
| Shi et al, 2021<sup>45</sup> | NR | No significant difference: IL-6 in 64 pts in therapy group in comparison to 35 pts in control group | Improvement in whole lung lesion volume from baseline to day 28 in MSC group (65 pts) compared with the control group (35 pts) | 28 |
| Lanzoni et al, 2021<sup>44</sup> | NR | IL-6 decreased in MSC group in comparison to control group (median) | NR | 31 |
| Shu et al, 2020<sup>37</sup> | Time of the oxygenation index to return to the normal range was faster in the MSC group, total 12 pts | Decreased IL-6, CRP in comparison to control; lymphocyte count return to normal faster after treatment in MSC group total 12 pts | Time for lung inflammation absorption was significantly shorter on CT imaging in the MSC group than in the control group. | 28 |
| Xu et al, 2021<sup>49</sup> | PaO<sub>2</sub>/ FiO<sub>2</sub> increase in 26 pts (mean) | No significant differences in CRP and IL-6 after treatment in 26 pts (mean) | Improved in 17 pts, no change in 3 pts among 20 pts of MSC group | 30 |
| Leng et al, 2020<sup>47</sup> | NR | Lymphocyte and WBC count increased in 1 pt, CRP decreased in 1 pt after treatment. | Improved in 1 pt of MSC group | 14 |
| Meng et al, 2020<sup>48</sup> | PaO<sub>2</sub>/ FiO<sub>2</sub> increased in 3 pts and decreased in 1 pt among MSC group | IL-6 decreased in 4 pts. CRP increased in 2 pts and decreased in 7 pts. D-dimer increased in 8 pts, decreased in 1 pt | Improved in 1 representative pt | 28 |
| Haberle et al, 2021<sup>46</sup> | PaO<sub>2</sub>/ FiO<sub>2</sub> increased in therapy group (5 pts) | Increased lymphocyte, decreased WBC count, CRP, IL-6 and no change of D-dimer in MSC group (5 pts) | NR | NR |
| Singh et al, 2020<sup>50</sup> | NR | WBC, IL-6 decreased in 4 pts and increased in 2 pts. Lymphocyte count increased in 3 pts and decreased in 3 pts. CRP increased in 1 pt and decreased in 5 pts | NR | 5 to 18 |
| Guo et al, 2020<sup>53</sup> | PaO<sub>2</sub>/ FiO<sub>2</sub> increased in 31 pts (median) | No change of WBC count in 31 pts (mean). Elevated lymphocyte count and decreased CRP, PCT, IL-6, D-dimer in 31 pts (median) | NR | NR |
| Chen et al, 2020<sup>51</sup> | NR | No change of IL-6, WBC, CRP, PCT in 25 pts (mean) | Improved in 16 pts<sup>#</sup> | NR |
| Peng et al, 2020<sup>58</sup> | Oxygenation index increased in 1 pt | Decreased lymphocyte count, CRP and IL-6 in 1 pt | Improved in 1 pt | 10 |
| Zhang et al, 2020<sup>67</sup> | NR | Decreased CRP, IL-6 and increased in lymphocyte count in 1 pt | Improved in 1 pt | 7 |
| Tang et al, 2020<sup>61</sup> | PaO<sub>2</sub>/ FiO<sub>2</sub> increased in 2 pts | WBC, IL-6 increased both in 1 pt and decreased in another 1 pt. Increased lymphocyte count in 2 pts, decreased CRP in 2 pts | NR | 12 & 14 |
| Zhu et al, 2020<sup>65</sup> | NR | Decreased CRP, PCT and D-dimer level in 1 pt. WBC and lymphocyte count increased in 1 pt | Improved in 1 pt | 14 |
| Zengin et al, 2020<sup>64</sup> | NR | Decreased CRP in 1 pt | Improved in 1 pt | 60 |
| Yilmaz R et al, 2020<sup>63</sup> | NR, Ejection fraction improved from 25% to 60% | Decreased CRP, PCT, D-dimer and increase in lymphocyte, WBC count in 1 pt | Improved in 1 pt | 26 |
| Tao et al, 2020<sup>62</sup> | PaO<sub>2</sub>/ FiO<sub>2</sub> decreased in 1 pt | Decreased D-dimer and CRP in 1 pt. Increased lymphocyte, WBC count and PCT in 1 pt | No improvement in 1 pt | 78 |
| Feng et al, 2020<sup>52</sup> | Oxygenation index increased in 8 pts (mean) | IL-6, CRP are in normal range after treatment. Increased lymphocyte and decrease WBC total count 16 pts (mean) | Improved in 16 pts | 28 |
| Liang et al, 2020<sup>56</sup> | NR | Decreased CRP, PCT, D-dimer, WBC and increased lymphocyte count in 1 pt | Improved in 1 pt | 17 |
| Rich et al, 2020<sup>66</sup> | NR | Decreased CRP and D-dimer in 1 pt. | Improved in 1 pt | 32 |
| Sahin et al, 2021<sup>59</sup> | NR | NR | NR | NR |
| Lu et al, 2021<sup>57</sup> | NR | Decreased WBC and IL-6 in 1 pt | Improved in 1 pt | 196 |
| Iglesias et al, 2021<sup>55</sup> | PaO<sub>2</sub>/ FiO<sub>2</sub> increased in 3 pts decreased in 2 pts | D-dimer increased in 3 pts and decreased in 2 patients. Lymphocyte count, PCT increased in 2 patients and decreased in 3 patients. CRP decreased in 5 pts | Improved in 3 pts | 21 |
| Hashemian et al 2021<sup>54</sup> | NR | IL-6 decreased in 5 pts and CRP decreased in 6 pts | Improved in 2 pts, no improvement in 1 pt | 60 |
| Sanchez-Guijo et al, 2020<sup>60</sup> | NR | Lymphocyte count increased in 5 pts among 6 pts. Decreased CRP in 8 pts among 9 pts and D-dimer in 5 pts among 8 pts | NR | 4 to 28 |

Aberrations: CRP, C-reaction protein; IL-6, interleukin-6; PCT, procalcitonin; WBC, white blood cell; NR, not reported; NA, not applicable; pts, patients

### Laboratory findings in patients

IL-6 level was reported in 20 patients with MSC cell therapy, of whom 17 (85%) reported a decrease. WBC count was reported in 14 patients with MSC cell therapy, with an increase in 7 (50%). D-dimer was reported in 24 MSC cell therapy patients with a decrease in 13 (54.2%) (Table 2).

### CT findings

CTs of the lungs were reported in 69 patients with MSC cell therapy, of whom improvement was reported in 64 (92.7%) patients, and no improvement/change in 5 (7.3%) patients (Table 2).

## DISCUSSION

This systematic review and meta-analysis evaluated 413 patients hospitalized for COVID-19. Of the 300 patients evaluated in 3 RCTs and 5 comparative observational studies, MSC cell administration was associated with significant reduction in all-cause mortality risk by 69%. MSC cell therapy was also associated with significant reduction of SAE risk by 64%. There was no significant difference in the occurrence of mild AEs. Of all patients who have received MSC cell therapy, pulmonary function was reported in 12 patients, showing improvement in 8 patients and worsening in 4 patients. IL-6 level was reported in only 20 patients, showing increase in 3 patients and decrease in 17 patients. Lung CTs showed improvement in 64 of the 69 patients reported.

Hospitalized COVID-19 patients are at significant risk of developing ARDS, multi-organ dysfunction syndrome, and acute respiratory failure that is associated with a poor prognosis. Those complications are thought to be the consequence of enhanced inflammation with inflammatory cytokine production and immune dysfunction triggered by SARS-CoV-2 infection.^68–70^ MSC cell therapy has been a focus of investigation because of its immunomodulatory effect that has been hypothesized to down regulate and suppress the inflammatory processes in COVID-19 patients.^16^

Safety is a critical issue for any new treatment, particularly in patients at high risk of death from the disease being managed. This analysis suggests a favorable safety profile with reduction of SAEs and no change in AEs in patients receiving MSC cell therapy. This safety profile is consistent with findings of other cell-based therapy trials targeting various pathologies.^71–74^ It is unlikely that these findings were affected by age differences between the treated and untreated groups, since the baseline ages were comparable (60.5 years of age in therapy groups and 60.4 years in control groups); this is important because older age is associated with increased rates of mortality from COVID-19.^75, 76^

Other outcomes examined included pulmonary function, clinical outcomes, and immune responses. Eighteen studies reported improvement of opacity in chest computed tomography within days of treatment. The improvement in pulmonary function and imaging findings also support further investigation of using MSC cell therapy for ARDS, especially in COVID-19 infected patients.

The findings from this meta-analysis also suggest beneficial effects of cell-based therapy on vital immunologic and inflammatory processes contributing to organ injury in SARS-CoV-2-infected patients. Seventeen of the studies reported reduction of inflammatory biomarkers after MSC cell therapy. MSC treatment appeared to mitigate the effects of cytokine release syndrome, a pathophysiological process in ARDS, and multi-organ dysfunction syndrome in severe cases of COVID-19. These findings suggest a potential mechanism of action and are consistent with previous results of preclinical and clinical studies for other diseases.^25^

Several important conclusions emerged from this review of the outcomes of MSC cell therapy for COVID-19. First, these data confirm previous findings of the low risks associated with MSC cell therapy. Our findings, combined with comparable studies treating various conditions provide strong support for MSC cell therapy having a favorable safety profile, even in this seriously ill population.^72–74, 77^ Second, MSC cell therapy was associated with markedly reduced mortality and SAEs in hospitalized COVID-19 patients. Among the patients included in this study, the risk of death in the group treated with MSC cell therapy was lowered by 69%. Third, although reported in limited studies, the biomarker findings support the hypothesis that MSC cell therapy reduces pathophysiologic and immunologic responses that contribute to death with SARS-CoV-2 infection.

To date, registry searches indicate that there are at least 112 clinical trials investigating the potential benefits of cell-based interventions in COVID-19 patients unresponsive to other available treatments. Nevertheless, additional larger trials are needed to further explore this promising therapeutic modality, given the limited success of other approaches to effectively manage severe cases of COVID-19. Other questions needing further study to be resolved include the best source of cells or cell products, especially for different stages of disease; the optimal dose and frequency of administration; whether the cells should be expanded in culture and, if so, for how long; and whether preconditioning the cells would improve their effectiveness. The results seen in this meta-analysis also suggest that MSC cell therapy may reduce the time to recovery and long-term complications from SARS-CoV-2 infection. Additional studies of cell-therapy in earlier phases of disease progression also are needed, especially to determine their potential ability to reduce the need for ICU admission, mechanical ventilation, and development of chronic inflammatory diseases.

### Limitations

Although this analysis found a reduction in mortality associated with MSC cell therapy, it is important to note that the conclusion was derived by comparing MSC cell therapy to the institutionally defined standard of care in the early phase of the COVID-19 pandemic. More recently, refinements of therapeutic approaches and vaccinations have considerably reduced mortality. The benefit of MSC cell therapy may be reduced or absent in patients not responding to these more contemporary treatments.

## CONCLUSIONS

This systematic meta-analysis demonstrates an association of MSC cell therapy with improvements in clinical outcomes, laboratory findings, and lung imaging in patients hospitalized for COVID-19, and a low incidence of adverse events related to treatment. The putative mechanisms of MSC cell therapy suggested by these and other clinical and pre-clinical studies, include beneficial modulation of inflammatory immune responses. While outcomes have improved with the changes in standard of care and vaccinations, patients continue to die from severe COVID-19. These findings support the urgent need for large, randomized double-blinded controlled trials to document the safety and efficacy of MSC cell therapy for the treatment and prevention of severe COVID-19. Appropriately designed studies would also help identify mechanisms that may potentially contribute to accelerate translation of cell-based therapies to other acute and chronic inflammatory diseases.

## Data Availability

Data analyzed in this study were a re-analysis of existing data, which are openly available at locations cited in the reference section.

## Acknowledgments

The authors wish to acknowledge Krishnendu Roy PhD from Georgia Institute of Technology for his expert opinion that was critical for this manuscript.

## Authors contribution

Wenchun Qu MD, MS, PhD: Conceptualization, Formal analysis, Methodology, Writing original draft, Writing review & editing, Final approval of the manuscript

Zhen Wang PhD: Conceptualization, Formal analysis, Methodology, Writing original draft, Writing review & editing, Final approval of the manuscript

Erica Engelberg-Cook DVM, PhD: Conceptualization, Writing original draft, Writing review & editing, Final approval of the manuscript

Abu Bakar Siddik MBBS: Conceptualization, Writing original draft, Writing review & editing, Final approval of the manuscript

Guojun Bu PhD: Conceptualization, Writing review & editing, Final approval of the manuscript

Julie G. Allickson PhD: Conceptualization, Writing review & editing, Final approval of the manuscript

Eva Kubrova MD: Conceptualization, Writing review & editing, Final approval of the manuscript

Arnold I. Caplan PhD: Conceptualization, Writing review & editing, Final approval of the manuscript

Joshua M. Hare MD: Conceptualization, Writing original draft, Writing review & editing, Final approval of the manuscript

Camillo Ricordi MD: Conceptualization, Writing original draft, Writing review & editing, Final approval of the manuscript

Carl J. Pepine MD: Conceptualization, Writing original draft, Writing review & editing, Final approval of the manuscript

Joanne Kurtzberg MD: Conceptualization, Writing original draft, Writing review & editing, Final approval of the manuscript, Final approval of the manuscript

Jorge M. Pascual MD: Conceptualization, Writing review & editing, Final approval of the manuscript

Jorge M. Mallea MD: Conceptualization, Writing review & editing, Final approval of the manuscript

Ricardo L. Rodriguez MD: Conceptualization, Writing review & editing, Final approval of the manuscript

Tarek Nayfeh MD: Conceptualization, Formal analysis, Writing review & editing, Final approval of the manuscript

Samer Saadi MD: Conceptualization, Writing review & editing, Final approval of the manuscript

Elaine M. Richards PhD Conceptualization, Writing review & editing, Final approval of the manuscript

Keith March MD, PhD : Conceptualization, Writing review & editing, Final approval of the manuscript

Fred P. Sanfilippo MD, PhD : Conceptualization, Writing review & editing, Final approval of the manuscript

---

**Table A1:** Inclusion and exclusion criteria.

| **PICOTS elements** | **Inclusion criteria** | **Exclusion criteria** |
| --- | --- | --- |
| Population | Patients with COVID-19 or SARS-CoV-2 infection | Animals |
| Interventions | Cell Therapy transplantation, include:<br>Stem cell<br>Mesenchymal stem cell<br>Mesenchymal stromal cell<br>Medicinal stem cells<br>MSC<br>iPS<br>iPSC<br>progenitor cell |  |
| Comparators | Usual care, supportive care only, no treatment | None |
| Outcomes | Adverse events, severe adverse events, mortality, inflammatory biomarker, PaO<sub>2</sub>/ FiO<sub>2</sub>, Imaging (CT), clinical general outcome | None |
| Timing | Any time | None |
| Settings | Setting (inpatient) | None |
| Study design | Randomized clinical trials, comparative observational studies and case reports/case series | In vitro studies, nonoriginal data (eg, narrative reviews, editorials, or erratum), cost-benefit analysis |
| Publications | Any language<br>Up to April 8, 2021 |  |

**Table A2:**
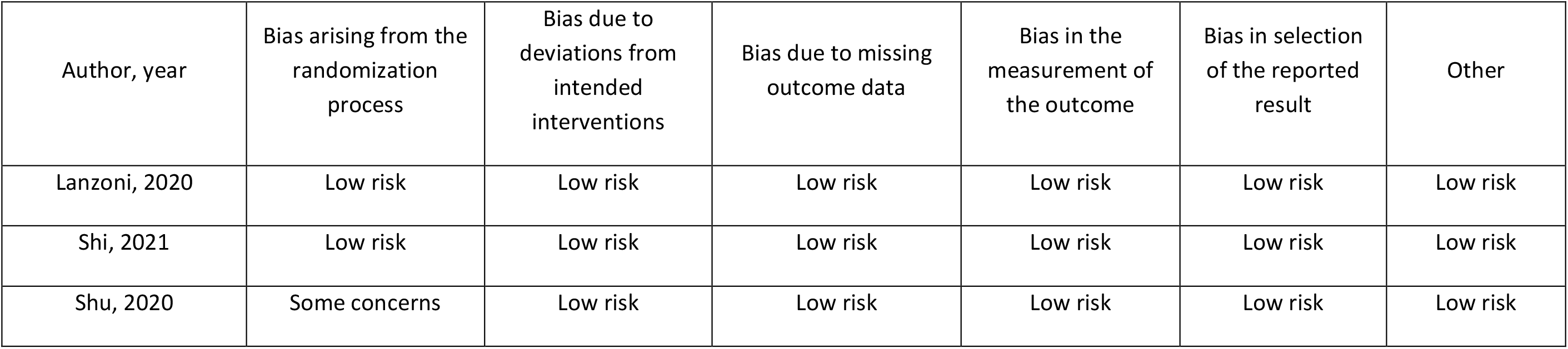
Risk of bias – Randomized Controlled Trials (RCTs)

| Author, year | Bias arising from the randomization process | Bias due to deviations from intended interventions | Bias due to missing outcome data | Bias in the measurement of the outcome | Bias in selection of the reported result | Other |
| --- | --- | --- | --- | --- | --- | --- |
| Lanzoni, 2020 | Low risk | Low risk | Low risk | Low risk | Low risk | Low risk |
| Shi, 2021 | Low risk | Low risk | Low risk | Low risk | Low risk | Low risk |
| Shu, 2020 | Some concerns | Low risk | Low risk | Low risk | Low risk | Low risk |

**Table A3:** Risk of bias – Comparative observational studies.

| Author, year | Selection |  |  |  | Comparability | Outcome |  |  |
| --- | --- | --- | --- | --- | --- | --- | --- | --- |
|  | Were the patients enrolled in the intervention group representative of the cohort 1? | Was the comparison group drawn from the same community as the intervention group? | Ascertainment of exposure by a secure record or structured interview? | Did the study demonstrate that the primary outcome was not present at the start of study? | Was the study controlled for baseline characteristics? | Was the primary outcome assessed by an independent blind assessor or record linkage? | Follow-up long enough for outcomes to occur? | Adequacy of follow-up |
| Xu, 2021 | Low risk | Low risk | Low risk | Yes | Yes | High risk | Low risk | Low risk |
| Haberle, 2021 | Low risk | Low risk | Low risk | Yes | Yes | High risk | Low risk | Low risk |
| Leng, 2020 | Low risk | Low risk | Low risk | Yes | Yes | Unclear | Low risk | Low risk |
|  | Were the patients enrolled in the intervention group representative of the cohort 1? | Was the comparison group drawn from the same community as the intervention group? | Ascertainment of exposure by a secure record or structured interview? | Did the study demonstrate that the primary outcome was not present at the start of study? | Was the study controlled for baseline characteristics? | Was the primary outcome assessed by an independent blind assessor or record linkage? | Follow-up long enough for outcomes to occur? | Adequacy of follow-up |
| Meng, 2020 | Low risk | Low risk | Low risk | Yes | Yes | High risk | Low risk | Low risk |
| Singh, 2020 | Low risk | Low risk | Low risk | Yes | Yes | High risk | Low risk | Low risk |

**Table A4:** Risk of bias – Non-comparative studies.

| Author,<br>Year | Selection | Ascertainment |  | Causality |  |  |  | Reporting |
| --- | --- | --- | --- | --- | --- | --- | --- | --- |
|  | Does the patient(s) represent(s) the whole experience of the investigator (center) or is the selection method unclear to the extent that other patients with similar presentation may not have been reported? | Was the exposure adequately ascertained? | Was the outcome adequately ascertained? | Were other alternative causes that may explain the observation ruled out? | Was there a challenge/rechallenge phenomenon? | Was there a dose–response effect? | Was follow-up long enough for outcomes to occur? | Is the case(s) described with sufficient details to allow other investigators to replicate the research or to allow practitioners make inferences related to their own practice? |
| Sahin, 2021 | Yes | Yes | Yes | No | No | No | Yes | No |
| Lu, 2021 | No | Yes | Yes | No | No | No | Yes | No |
| Iglesias, 2021 | Yes | Yes | Yes | No | No | No | Yes | Yes |
| Hashemian, 2021 | Yes | Yes | Yes | No | No | No | Yes | Yes |
| Liang, 2020 | No | Yes | Yes | No | No | No | Yes | No |
| Peng, 2020 | No | Yes | Yes | No | No | No | Yes | No |
|  | Does the patient(s) represent(s) the whole experience of the investigator (center) or is the selection method unclear to the extent that other patients with similar presentation may not have been reported? | Was the exposure adequately ascertained? | Was the outcome adequately ascertained? | Were other alternative causes that may explain the observation ruled out? | Was there a challenge/rechallenge phenomenon? | Was there a dose–response effect? | Was follow-up long enough for outcomes to occur? | Is the case(s) described with sufficient details to allow other investigators to replicate the research or to allow practitioners make inferences related to their own practice? |
| Rich, 2020 | Yes | Yes | Yes | No | No | No | Yes | No |
| Tang, 2020 | No | Yes | Yes | No | No | No | Yes | No |
| Tao, 2020 | No | Yes | Yes | No | No | No | Yes | No |
| Yilmaz, 2020 | No | Yes | Yes | No | No | No | Yes | No |
| Zengin, 2020 | No | Yes | Yes | No | No | No | Yes | No |
| Zheng, 2020 | No | Yes | Yes | No | No | No | Yes | No |
| Zhu, 2020 | No | Yes | Yes | No | No | No | Yes | No |
|  | Does the patient(s) represent(s) the whole experience of the investigator (center) or is the selection method unclear to the extent that other patients with similar presentation may not have been reported? | Was the exposure adequately ascertained? | Was the outcome adequately ascertained? | Were other alternative causes that may explain the observation ruled out? | Was there a challenge/rechallenge phenomenon? | Was there a dose–response effect? | Was follow-up long enough for outcomes to occur? | Is the case(s) described with sufficient details to allow other investigators to replicate the research or to allow practitioners make inferences related to their own practice? |
| Chen, 2020 | Yes | Yes | Yes | No | No | No | Yes | Yes |
| Sanchez-Guijo, 2020 | Yes | Yes | Yes | No | No | No | Yes | Yes |
| Feng, 2020 | Yes | Yes | Yes | No | No | No | Yes | Yes |
| Guo, 2020 | Yes | Yes | Yes | No | No | No | Yes | Yes |

**TABLE A5:** Severe adverse events in cell therapy group.

| **Events** | **Numbers of events** |
| --- | --- |
| Multi-organ failure | 5 |
| Shock | 4 |
| GI bleeding | 3 |
| Liver dysfunction | 2 |
| Fungal pneumonia | 2 |
| Bacterial pneumonia | 2 |
| ARDS | 1 |
| Pneumothorax | 1 |
| Abnormal blood clotting | 1 |
| Myocardial damage | 1 |
| Expiratory dyspnea | 1 |
| Heart failure | 1 |
| Seizure | 1 |
| Cardiac arrest | 1 |
| Multi-organ failure/sepsis | 1 |
| AKI | 1 |
| Pulmonary edema | 1 |
| Bilateral pleural effusion | 1 |
| Bacteremia | 1 |
| Not specified | 2 |

Abbreviation: ARDS, acute respiratory distress syndrome; AKI, acute kidney injury

**FIGURE A1:**
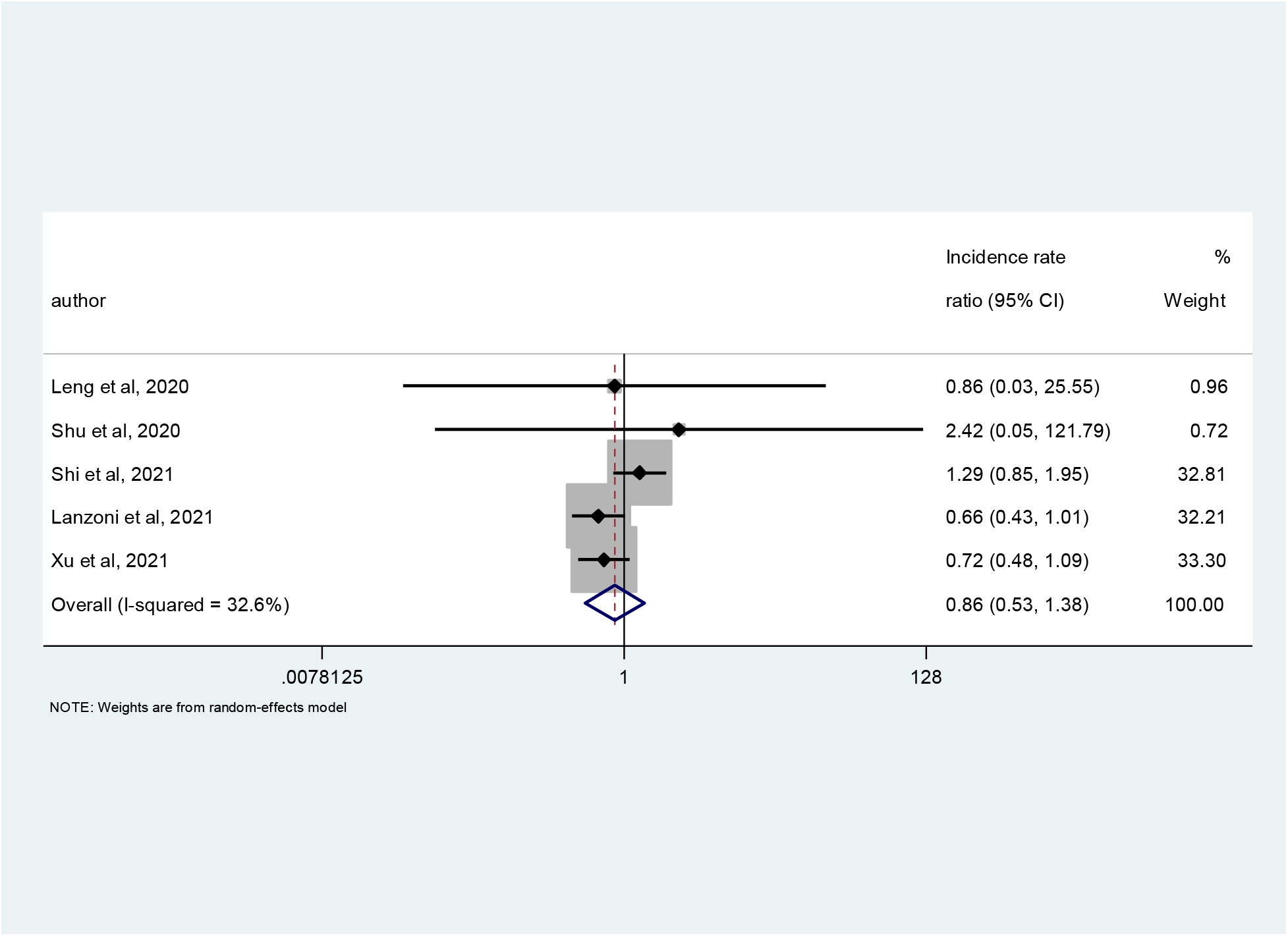
Adverse events.

Database(s): EBM Reviews - Cochrane Central Register of Controlled Trials March 2021, EBM Reviews - Cochrane Database of Systematic Reviews 2005 to April 8, 2021, Embase 1974 to 2021 April 08, Ovid MEDLINE(R) and Epub Ahead of Print, In-Process, In-Data-Review & Other Non-Indexed Citations and Daily 1946 to April 08, 2021 Search Strategy:

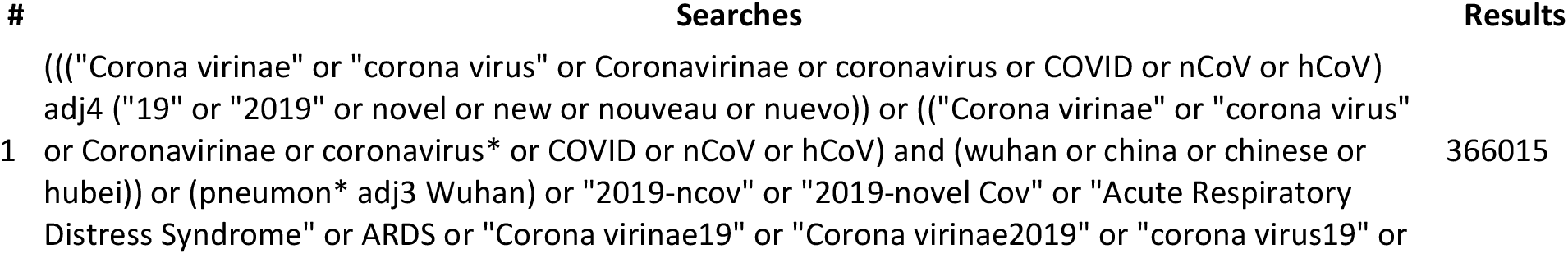
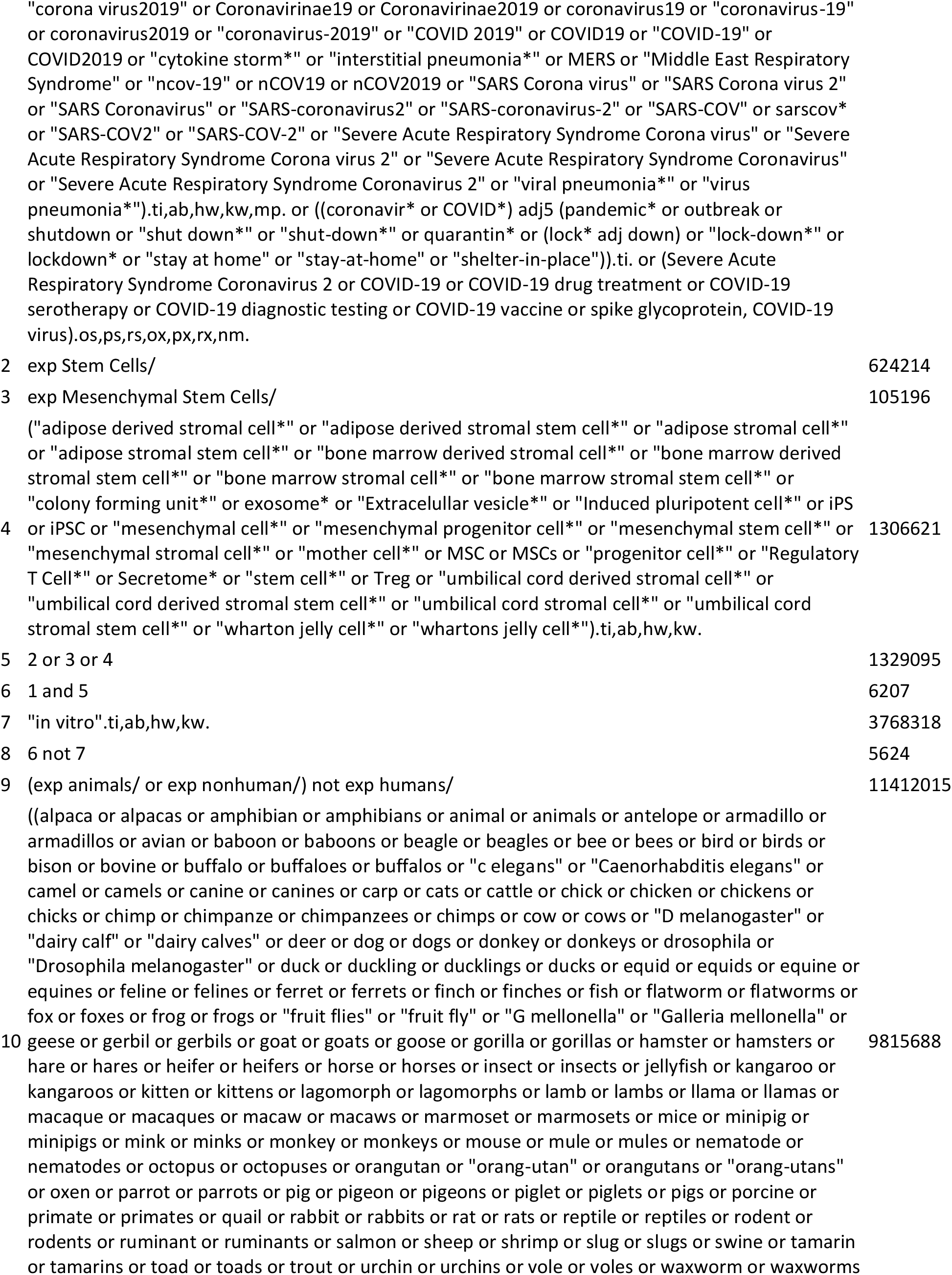
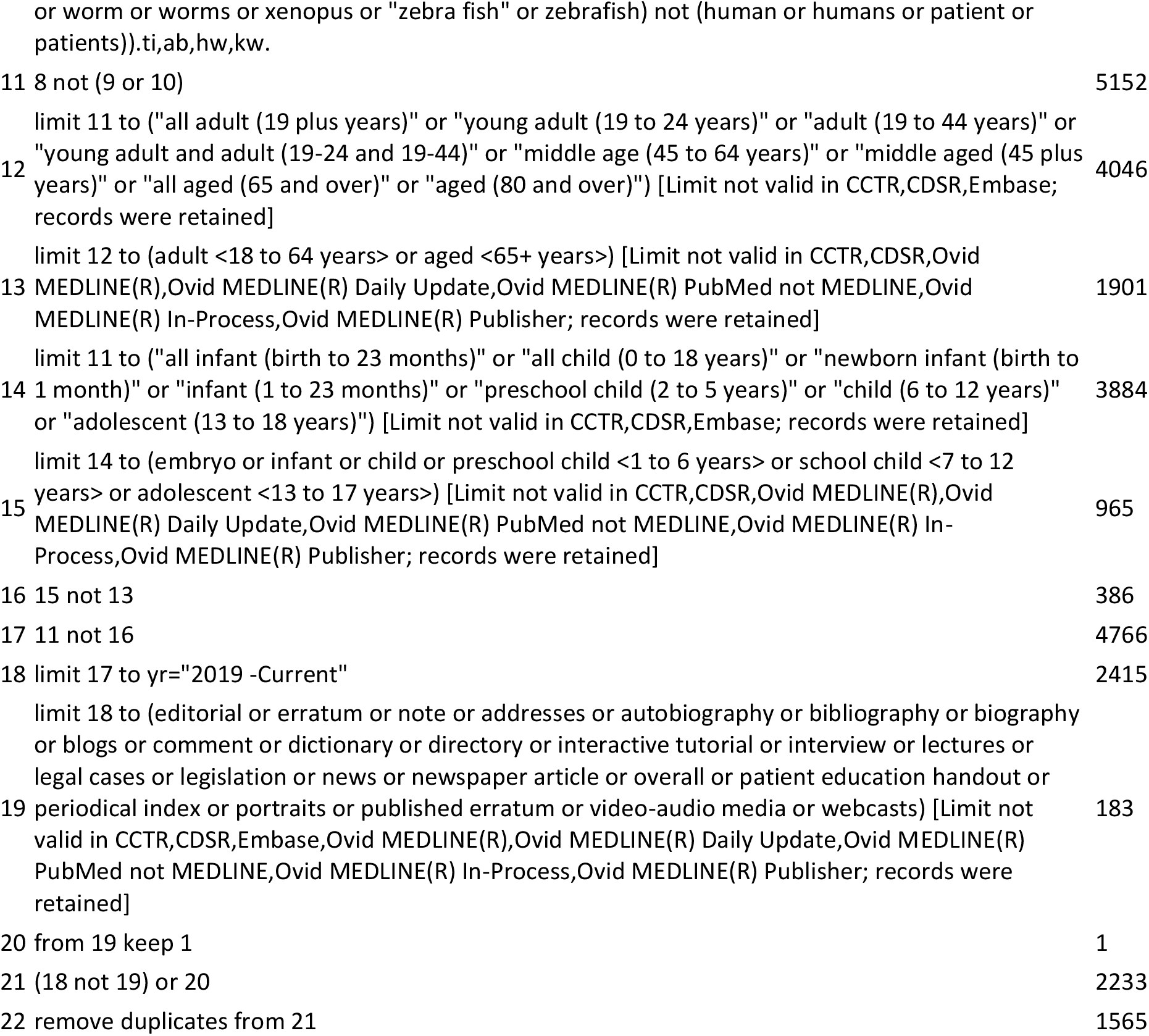

## Scopus

1. TITLE-ABS-KEY(((“Corona virinae” or “corona virus” or Coronavirinae or coronavirus or COVID or nCoV or hCoV) W/4 (“19” or “2019” or novel or new or nouveau or nuevo)) OR ((“Corona virinae” or “corona virus” or Coronavirinae or coronavirus* or COVID or nCoV or hCoV) and (wuhan or china or chinese or hubei)) OR (pneumon* W/3 Wuhan) OR “2019-ncov” OR “2019-novel Cov” OR “Acute Respiratory Distress Syndrome” OR ARDS OR “Corona virinae19” OR “Corona virinae2019” OR “corona virus19” OR “corona virus2019” OR Coronavirinae19 OR Coronavirinae2019 OR coronavirus19 OR “coronavirus-19” OR coronavirus2019 OR “coronavirus-2019” OR “COVID 2019” OR COVID19 OR “COVID-19” OR COVID2019 OR “cytokine storm*” OR “interstitial pneumonia*” OR MERS OR “Middle East Respiratory Syndrome” OR “ncov-19” OR nCOV19 OR nCOV2019 OR “SARS Corona virus” OR “SARS Corona virus 2” OR “SARS Coronavirus” OR “SARS-coronavirus2” OR “SARS-coronavirus-2” OR “SARS-COV” OR sarscov* OR “SARS-COV2” OR “SARS-COV-2” OR “Severe Acute Respiratory Syndrome Corona virus” OR “Severe Acute Respiratory Syndrome Corona virus 2” OR “Severe Acute Respiratory Syndrome Coronavirus” OR “Severe Acute Respiratory Syndrome Coronavirus 2” OR “viral pneumonia*” OR “virus pneumonia*”)
2. TITLE(((coronavir* or COVID*) W/5 (pandemic* or outbreak or shutdown or “shut down*” or “shut-down*” or quarantin* or (lock* W/1 down) or “lock-down*” or lockdown* or “stay at home” or “stay-at-home” or “shelter-in-place”)))
3. TITLE-ABS-KEY(“adipose derived stromal cell*” OR “adipose derived stromal stem cell*” OR “adipose stromal cell*” OR “adipose stromal stem cell*” OR “bone marrow derived stromal cell*” OR “bone marrow derived stromal stem cell*” OR “bone marrow stromal cell*” OR “bone marrow stromal stem cell*” OR “colony forming unit*” OR exosome* OR “Extracelullar vesicle*” OR “Induced pluripotent cell*” OR iPS OR iPSC OR “mesenchymal cell*” OR “mesenchymal progenitor cell*” OR “mesenchymal stem cell*” OR “mesenchymal stromal cell*” OR “mother cell*” OR MSC OR MSCs OR “progenitor cell*” OR “Regulatory T Cell*” OR Secretome* OR “stem cell*” OR Treg OR “umbilical cord derived stromal cell*” OR “umbilical cord derived stromal stem cell*” OR “umbilical cord stromal cell*” OR “umbilical cord stromal stem cell*” OR “wharton jelly cell*” OR “whartons jelly cell*”)
4. PUBYEAR AFT 2018
5. (1 or 2) and 3 and 4
6. TITLE-ABS-KEY(“in vitro”)
7. 5 and not 6
8. TITLE-ABS-KEY((alpaca OR alpacas OR amphibian OR amphibians OR animal OR animals OR antelope OR armadillo OR armadillos OR avian OR baboon OR baboons OR beagle OR beagles OR bee OR bees OR bird OR birds OR bison OR bovine OR buffalo OR buffaloes OR buffalos OR “c elegans” OR “Caenorhabditis elegans” OR camel OR camels OR canine OR canines OR carp OR cats OR cattle OR chick OR chicken OR chickens OR chicks OR chimp OR chimpanze OR chimpanzees OR chimps OR cow OR cows OR “D melanogaster” OR “dairy calf” OR “dairy calves” OR deer OR dog OR dogs OR donkey OR donkeys OR drosophila OR “Drosophila melanogaster” OR duck OR duckling OR ducklings OR ducks OR equid OR equids OR equine OR equines OR feline OR felines OR ferret OR ferrets OR finch OR finches OR fish OR flatworm OR flatworms OR fox OR foxes OR frog OR frogs OR “fruit flies” OR “fruit fly” OR “G mellonella” OR “Galleria mellonella” OR geese OR gerbil OR gerbils OR goat OR goats OR goose OR gorilla OR gorillas OR hamster OR hamsters OR hare OR hares OR heifer OR heifers OR horse OR horses OR insect OR insects OR jellyfish OR kangaroo OR kangaroos OR kitten OR kittens OR lagomorph OR lagomorphs OR lamb OR lambs OR llama OR llamas OR macaque OR macaques OR macaw OR macaws OR marmoset OR marmosets OR mice OR minipig OR minipigs OR mink OR minks OR monkey OR monkeys OR mouse OR mule OR mules OR nematode OR nematodes OR octopus OR octopuses OR orangutan OR “orang-utan” OR orangutans OR “orang-utans” OR oxen OR parrot OR parrots OR pig OR pigeon OR pigeons OR piglet OR piglets OR pigs OR porcine OR primate OR primates OR quail OR rabbit OR rabbits OR rat OR rats OR reptile OR reptiles OR rodent OR rodents OR ruminant OR ruminants OR salmon OR sheep OR shrimp OR slug OR slugs OR swine OR tamarin OR tamarins OR toad OR toads OR trout OR urchin OR urchins OR vole OR voles OR waxworm OR waxworms OR worm OR worms OR xenopus OR “zebra fish” OR zebrafish) AND NOT (human OR humans or patient or patients))
9. 7 and not 8
10. TITLE-ABS-KEY(newborn* or neonat* or infant* or toddler* or child* or adolescent* or paediatric* or pediatric* or girl or girls or boy or boys or teen or teens or teenager* or preschooler* or “pre-schooler*” or preteen or preteens or “pre-teen” or “pre-teens” or youth or youths) AND NOT TITLE-ABS-KEY(adult or adults or “middle age” or “middle aged” OR elderly OR geriatric* OR “old people” OR “old person*” OR “older people” OR “older person*” OR “very old”)
11. 9 and not 10
12. DOCTYPE(ed) OR DOCTYPE(bk) OR DOCTYPE(er) OR DOCTYPE(no) OR DOCTYPE(sh)
13. 11 and not 12
14. INDEX(embase) OR INDEX(medline) OR PMID(0* OR 1* OR 2* OR 3* OR 4* OR 5* OR 6* OR 7* OR 8* OR 9*)
15. 13 and not 14

## ClinicalTrials.Gov

Condition or disease

(“2019 novel coronavirus” OR “2019-nCoV” OR “acute respiratory distress” OR “acute respiratory syndrome” OR ARDS OR Coronavirus OR “COVID 19” OR “cytokine storm” OR “interstitial pneumonia” OR MERS OR “Middle East Respiratory Syndrome” OR “SARS-CoV”)

(“viral pneumonia” OR “virus pneumonia”)

Other terms

(“colony forming unit*” OR exosome* OR “Extracelullar vesicle*” OR “Induced pluripotent cell*” OR iPS OR iPSC OR “mesenchymal cell*” OR “mesenchymal progenitor cell*” OR “mother cell*” OR MSC OR MSCs OR “progenitor cell*” OR “Regulatory T Cell*”)

(Secretome* OR “stem cell*” OR “stromal cell*” OR Treg OR “wharton jelly cell*” OR “whartons jelly cell*”) Limited to Adults

First posting

01/01/2019 To 01/12/2021

